# A rapid and simple protocol for concentration of SARS-CoV-2 from sewage

**DOI:** 10.1101/2021.05.27.21257934

**Authors:** Diana P. Wehrendt, Mariana G. Massó, Adrián Gonzales Machuca, Claudia V. Vargas, Melina E. Barrios, Josefina Campos, Damián Costamagna, Luis Bruzzone, Daniel M. Cisterna, Néstor Gabriel Iglesias, Viviana A. Mbayed, Elsa Baumeister, Daniela Centrón, María Paula Quiroga, Leonardo Erijman

**Affiliations:** Instituto de Investigaciones en Ingeniería Genética y Biología Molecular “Dr Héctor N. Torres” (INGEBI-CONICET) Vuelta de Obligado 2490 - C1428ADN; Buenos Aires, Argentina; Consejo Nacional de Investigaciones Científicas y Técnicas (CONICET); Laboratorio de Investigaciones en Mecanismos de Resistencia a Antibióticos. Instituto de Investigaciones en Microbiología y Parasitología Médica, Facultad de Medicina, Universidad de Buenos Aires (IMPaM, UBA-CONICET); Instituto de Investigaciones en Producción Animal (INPA); Facultad de Farmacia y Bioquimica. Universidad de Buenos Aires; Instituto de Investigaciones en Bacteriología y Virologia Molecular (IBAViM); Instituto Nacional de Enfermedades Infecciosas (INEI) ANLIS “Dr Carlos G. Malbrán”; Autoridad del Agua. Gobierno de la Provincia de Buenos Aires; Aguas Bonaerenses SA; Laboratorio de Virus Emergentes, Departamento de Ciencia y Tecnología, Universidad Nacional de Quilmes; Servicio Virosis Respiratorias, Laboratorio Nacional de Referencia de Enfermedades Respiratorias Virales, INEI-ANLIS “Dr Carlos G Malbrán”. Centro Nacional de Influenza de OMS; Departamento de Fisiología, Biología Molecular y Celular, Facultad de Ciencias Exactas y Naturales. Universidad de Buenos Aires

**Keywords:** Environmental surveillance, SARS-CoV-2, Sewage, Virus concentration, Wastewater based epidemiology

## Abstract

The aim of this study was to set up a simple protocol to concentrate SARS-CoV-2 from sewage, which can be implemented in laboratories with minimal equipment resources. The method avoids the need for extensive purification steps and reduces the concentration of potential inhibitors of RT-qPCR contained in sewage. The concentration method consists of a single step, in which a small volume of sewage sample is incubated with polyaluminum chloride (PAC). Virus particles adsorbed to the precipitate are collected by low-speed centrifugation, after which the recovered pellet is resuspended with a saline buffer. The PAC concentration method produced an average shift of 4.4-units in Cq values compared to non-concentrated samples, indicating a 25-fold increase in detection sensitivity. The lower detection limit corresponded approximately to 100 copies per ml. Kappa index indicated substantial agreement between PAC and PEG precipitation protocols (k=0.688, CI 0.457-0.919). PAC concentrated samples can be processed immediately for RNA purification and qPCR or sent refrigerated to a diagnosis center, where SARS-CoV-2 detection should be performed in the same way as for clinical samples. This low cost protocol could be useful to aid in the monitoring of community circulation of SARS-CoV-2, especially in low- and middle-income countries, which do not have massive access to support from specialized labs for sewage surveillance.

## Main text

One of the main challenges faced by health authorities during the Covid-19 pandemics has been testing for SARS-CoV-2 on a large enough scale. It has been shown that analysis of sewage can complement clinical testing by providing a fair representation of the incidence of SARS-CoV-2 within a community, including pre-symptomatic and asymptomatic individuals (Foladori et al., 2020). For these reasons, SARS-CoV-2 sewage surveillance has the potential to provide early warning of the emergence of new outbreaks. Sewage screening may also be a useful tool in evaluating the efficacy of the incipient vaccination campaign.

Methods for the detection of SARS-CoV-2 (RNA extraction and RT-qPCR detection) in sewage samples are the same as those used for testing nasopharyngeal samples. However, viral particles in sewage are usually very diluted and therefore require a concentration step before RNA extraction. To be considered effective and applicable, the concentration method has to provide highly efficient viral recovery and must be repeatable within a laboratory, as well as reproducible between laboratories. There is no single standardized method for the concentration of SARS-CoV-2 in sewage. Instead, there are a wide variety of techniques to recover SARS-CoV-2 from wastewater used by different laboratories around the world. The most commonly used methods to successfully concentrate viral particles from sewage include adsorption-elution with negatively charged membrane, ultrafiltration with centrifugal concentrators, PEG-precipitation and ultracentrifugation (Ahmed et al., 2020; Philo et al., 2021). These procedures require instrumentation that is not always readily available in small laboratories, including those in local municipal wastewater treatment plants. This restricts the possibility of many villages and small towns to gain the information on virus circulation in the community that is afforded by sewage surveillance.

Viruses can also be concentrated from wastewater by aluminum-driven flocculation. Aluminum hydroxide adsorption-precipitation is a general method used to concentrate viruses from water, wastewater, and eluates from adsorbent filters (AWWA, 2018). The technique shows acceptable efficiency recovery for SARS-CoV-2 (Barril et al., 2021; Randazzo et al., 2020), though it requires extensive purification procedures, especially for sewage samples containing high amounts of suspended solids. This is a drawback because the solid component of sewage may contain a higher number of SARS-CoV-2 RNA copies than the corresponding liquid phase (Graham et al., 2021; Michael-Kordatou et al., 2020).

We describe here a simple method, applicable to sewage within an ample range of Biochemical Oxygen Demand (BOD) strength, which avoids the need for extensive purification steps and reduces the concentration of potential inhibitors of RT-qPCR contained in sewage. The concentration protocol is simple enough to be performed in any low-resource laboratory. The concentrated sample can be sent to a microbiology laboratory, where it should be handled as any clinical sample for RNA extraction and SARS-CoV-2 RT-qPCR detection.

The concentration method consisted of a single step, in which a small volume of sewage sample is incubated with poly-aluminum chloride (PAC). This inorganic polymer has been used worldwide in water and wastewater treatment for decades as a primary coagulant aid for clarification and phosphorus removal. Total aluminum content of PAC products, expressed as Al2O3, ranges from about 6 to 24% by weight in aqueous solutions. The mechanisms of adsorption appear to involve electrostatic interactions between the negatively charged virus surface and the positively charged aluminum hydroxide surfaces and coordination of the virus surface by hydroxo-aluminum complexes (AWWA, 2018). Virus particles adsorbed to the precipitate are collected by low-speed centrifugation, after which the recovered pellet is resuspended with a saline buffer (Figure 1). The concentrated samples are stable for at least 1 week at 4°C (Table S1), and therefore can be processed immediately for RNA purification and RT-qPCR or sent refrigerated to a diagnosis center, where SARS-CoV-2 should be tested in the same way as on a clinical sample.

**Figure 1:**
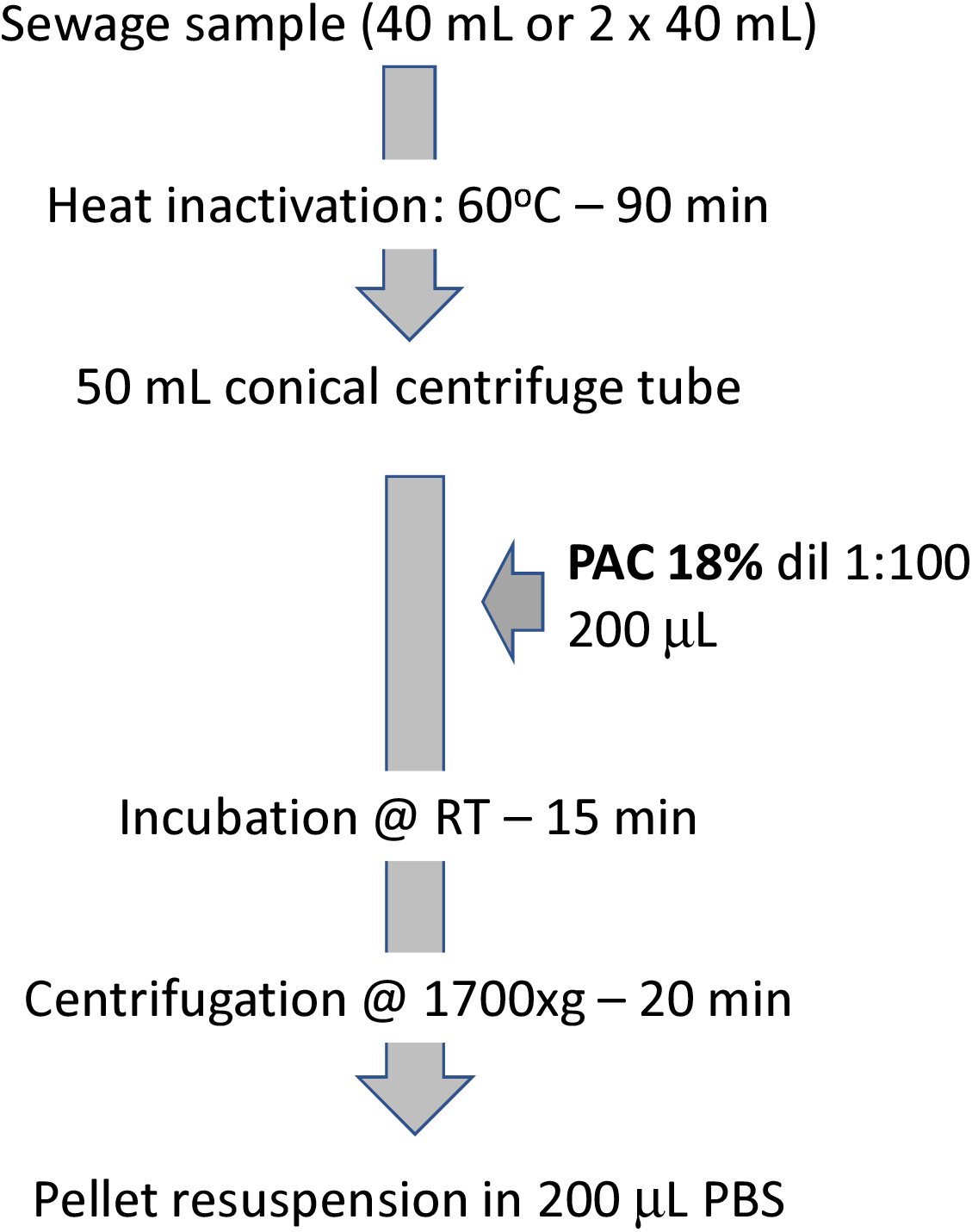
Simplified method of SARS-CoV-2 RNA extraction from sewage samples

A total of 60 sewage samples were collected weekly during a 15-weeks period at pumping stations of two wastewater treatment plants located in the Buenos Aires Metropolitan Area (AK, 20,000 population equivalents (pe) and SV, 15,000 pe), and at sewer manholes of two apartment block complexes (SP, 3500 pe and SF, 5000 pe). Biological oxygen demand of collected samples ranged from 248 ± 33 mg/L in SF to 630 ± 116 mg/L in SP. Grab samples (250 mL) were transported within 3h of collection and processed immediately upon arrival to the laboratory. For the purpose of operator protection, samples were initially incubated at 60°C during 90 minutes for heat inactivation of the virus. Then, 200μl of a 1/100 PAC solution were added to 40 ml of pre-inactivated wastewater samples (1/20,000 PAC final dilution). The pH was adjusted, if necessary, within the range of 6-7. Incubation was carried out for 15 min with agitation at room temperature. Samples were centrifuged at 1,700xg for 20 min at room temperature and the resultant pellet was resuspended in 200μl of phosphate buffer saline (PBS). RNA purification was performed using the Viral Nucleic Extraction Kit II (Geneaid), following manufacturer instructions, with the addition of a short spin after the lysis step, to prevent column clogging. RNA was eluted in 50µL of RNAse-free water.. SARS-CoV-2 was detected using the DisCoVery SARS-CoV-2 RT-PCR detection kit (APBiotech), using 5µL of purified RNA in a final reaction volume of 25µL. The targets of this kit are SARS-CoV-2 nucleocapsid (N) and orf1 genes. Human RNAseP was used as an internal control.

The concentration method produced a shift of an average of 4.4±1.9 in Cq values compared to non-concentrated samples (n=24, Fig 2), indicating a 25-fold increase in detection sensitivity. To estimate the detection limit of the method, a 40mL sewage sample taken during the early stage of the Covid-19 pandemics, i.e. negative for SARS-CoV-2, was artificially seeded with a 10-µL droplet of an inactivated virus culture (2.33×10^5^ tissue culture infectious dose TCID50/mL, corresponding to approximately 4×10^9^ copies/μL). The inoculated sample was 10-fold serially diluted in sewage samples negative for SARS-CoV-2. Each dilution was subsequently subjected to the concentration protocol followed by RNA extraction and qPCR detection. The lower detection limit was the 10^−7^ dilution, corresponding to an initial viral genome concentration of approximately 100 copies /mL.

**Figure 2:**
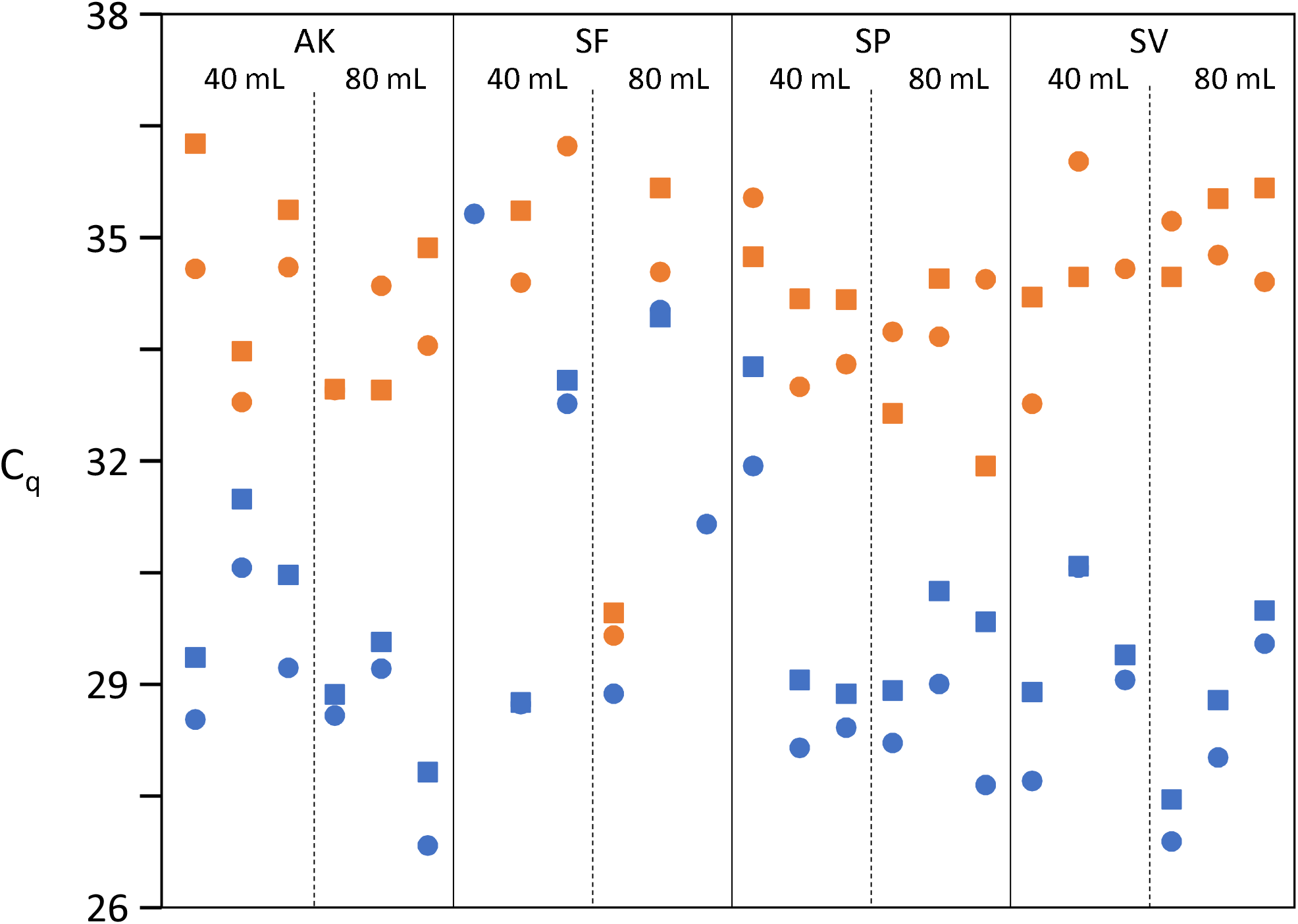
RT-qPCR charts for each treatment plant before (orange) and after PAC concentration (light blue). Cq above 40 was considered a non-detection. Circles and squares represent the targets of the RT-qPCR, respectively SARS-CoV-2 nucleocapsid (N) and orf1 genes.

By starting with a low initial volume of raw sewage (40 mL or 80 mL), we were able to perform RNA extraction directly from the pellet obtained after precipitation, and therefore the use of beef extract in the final purification step (AWWA, 2018) could be omitted. Nevertheless, because PAC is a nonspecific coagulant, other substances such as humic substances contained in the sewage (Mendoza et al., 2020) may still be concentrated with the virus. The presence of such impurities may not be removed in the RNA extraction step and might cause the concentrated sample to be inhibitory to the RT-qPCR assay. Therefore, although virus precipitation could be potentially increased by using larger amounts of Al(OH)3 and larger sample volumes, the optimum values of sample volume and PAC concentration was a rather delicate balance for maximum virus recovery with minimal enzyme inhibition. We note that the physical and chemical properties of the numerous PAC products can vary, including their content of other inorganic salts, which can affect its properties. Therefore, it is recommended to validate the optimum PAC concentration for each different batch of commercial PAC.

Covid-19 incidence in the metropolitan area that includes the catchment area of the wastewater treatment plants showed a significantly negative correlation with the Cq of SARS-CoV-2 detection (Spearman’s correlation r =-0.635, p= 0.00463). Despite the differences in sewage characteristics, the method could consistently detect the presence of SARS-CoV-2 in the four samples, even under relatively low Covid-19 incidence (<45 reported active cases / 100,000 people). Importantly, we did not detect major changes in Cq when concentrated samples were stored at 4°C in PBS for at least one week before RNA extraction (Table S1).

A kappa index (Cohen, 1960) was calculated to measure the agreement between tests performed after the PAC concentration protocol and a PEG/NaCl concentration method (Wu et al., 2020), which had been run in parallel on the same samples. The result was kappa = 0.688; SE = 0.118; 95% confidence interval 0.457 to 0.919; n=60), indicating substantial agreement between the two concentration methods.

In conclusion, we describe here a rapid and simple protocol that can efficiently concentrate SARS-CoV-2 from sewage, which can be straightforwardly performed in a laboratory with low infrastructure. Importantly, our data indicate that this low-cost method is suitable to concentrate efficiently SARS-CoV-2 from sewage with different levels of settleable solids. Concentrated samples can be sent refrigerated to a diagnosis center, where they may be handled as any other sample received for SARS-CoV-2 detection. This protocol could be useful to aid in the monitoring of community circulation of SARS-CoV-2, especially in low- and middle-income countries, which do not have massive access to support from specialized labs for sewage surveillance.

## Supporting information

Effect of storage during 7 days at 4oC of concentrated sewage samples on RT-qPCR detection of SARS-CoV-2

## Data Availability

All data will be available upon request

## Acknowledgments

This work was funded by the Programa de Articulación y Fortalecimiento Federal de las Capacidades en Ciencia y Tecnología COVID-19 (Proyecto BSAS-32; Ministerio de Ciencia, Tecnología e Innovación de la Nación (MINCyT) to LE and by Grant IP No. 13 COVID 19 from ANPCyT to EB. We thank all members of the network for the detection of Coronavirus in the Environment (MINCyT) for helpful discussions. We appreciate the essential collaboration of Aguas Bonaerenses SA (ABSA), Autoridad del Agua (ADA) and Organismo Provincial para el Desarrollo Sostenible (OPDS). Carlos Zapata and Matias Goñi are gratefully acknowledged for the collection and transportation of samples.

