## Supplementary material for "A rapid and simple protocol for concentration of SARS-CoV-2 from sewage": Effect of storage during 7 days at 4oC of concentrated sewage samples on RT-qPCR detection of SARS-CoV-2

### Supplemental Material

Table S1: Effect of storage during 7 days at 4°C of concentrated sewage samples on RT-qPCR detection of SARS-CoV-2

| Sample ID | Target gene | | | |
|---|---|---|---|---|
| | N | | Orf1ab | |
| | Before storage | After storage | Before storage | After storage |
| AK31 | 31,18 | 33,24 | 32,52 | 38,89 |
| AK32 | 31,33 | 33,27 | 32,10 | 36,72 |
| SF29 | 30,84 | 30,59 | 31,24 | 30,80 |
| SF31 | 35,37 | 34,1 | 35,85 | n.d. |
| SF32 | 34,52 | n.d. | 44,26 | 37,67 |
| SP29 | 31,87 | 30,5 | 33,88 | 31,48 |
| SP32 | 35,94 | 35,2 | n.d. | n.d. |
| SV31 | 33,63 | 33,06 | n.d. | 34,23 |
| SV32 | 29,76 | 30,3 | 31,05 | 31,91 |

n.d.: non- detection.
